# COVID-19, smoking, and inequalities: a cross-sectional survey of adults in the UK

**DOI:** 10.1101/2020.04.30.20086074

**Authors:** Sarah E. Jackson, Jamie Brown, Lion Shahab, Andrew Steptoe, Daisy Fancourt

## Abstract

**Objectives:** To examine associations between smoking and COVID-19 relevant outcomes, taking into account the influence of inequalities and adjusting for potential confounding variables.

**Design:** Online cross-sectional survey.

**Setting:** UK.

**Participants:** 53,002 men and women aged ≥18y.

**Main outcome measures:** Confirmed and suspected COVID-19, worry about catching and becoming seriously ill from COVID-19, and adherence to protective behaviours. Socioeconomic position was defined according to highest level of education (post-16 qualifications: yes/no).

**Results:** Compared with never smokers (0.3% [95%CI 0.2-0.3%]), prevalence of confirmed COVID-19 was higher among current (0.6% [0.4-0.8%]) but not ex-smokers (0.2% [0.2-0.3%]). The associations were similar before (current: OR 2.14 [1.49-3.08]; ex-smokers: OR 0.73 [0.47-1.14]) and after (current: OR 1.79 [1.22-2.62]; ex-smokers: OR 0.85 [0.54-1.33]) adjustment for potential confounders. For current smokers, this was moderated by socioeconomic position, with higher rates relative to never smokers only seen in those without post-16 qualifications (OR 3.53 [2.04-6.10]). After including suspected cases, prevalence was higher among current smokers (11.2% [10.6-11.9%], OR 1.11 [1.03-1.20]) and ex-smokers (10.9% [10.4-11.5%], OR 1.07 [1.01-1.15]) than never smokers (10.2% [9.9-10.6%]), but remained higher only among ex-smokers after adjustment (OR 1.21 [1.13-1.29]). Current and ex-smokers had higher odds than never smokers of reporting significant stress about catching (current: OR 1.43 [1.35-1.52]; ex-smokers: OR 1.15 [1.09-1.22]) or becoming seriously ill from COVID-19 (current: OR 1.34 [1.27-1.43]; ex-smokers: OR 1.22 [1.16-1.28]). Adherence to recommendations to prevent the spread of COVID-19 was generally high (96.3% [96.1-96.4%]), but lower among current than never smokers (OR 0.70 [0.62-0.78]).

**Conclusions:** When assessed by self-report in a population sample, current smoking was independently associated with confirmed COVID-19 infection. There were socioeconomic disparities, with the association only apparent among those without post-16 qualifications. Smokers reported lower adherence to guidelines despite being more worried than non-smokers about catching or becoming seriously ill from COVID-19.

**Registration:** The analysis plan was pre-registered on Open Science Framework (https://osf.io/pcs49/).

*What is already known on this topic:*

- Former or current smoking can increase the risk of respiratory viral and bacterial infections and is associated with worse outcomes for those infected.
- However, data from several countries indicate that rates of current smoking are substantially lower among hospitalised COVID-19 patients than would be expected based on population-level smoking prevalence.

*What this study adds:*

- Data from a large population-based sample of adults in the UK conflict with the hypothesis that smoking is protective against COVID-19 infection; rather, we found that current smoking was independently associated with increased odds of confirmed COVID-19 infection after adjusting for relevant confounders.
- Socioeconomic disparities were evident, with the association between smoking and confirmed COVID-19 only apparent among those without post-16 qualifications.
- Smokers reported lower adherence to guidelines despite being more worried than non-smokers about catching or becoming seriously ill from COVID-19.

## Introduction

The influence of smoking on COVID-19 infection and outcomes is unclear. More than a billion people around the world smoke tobacco, and the vast majority live in low- and middle-income countries or belong to more disadvantaged socioeconomic groups (1,2). Early data have not provided clear evidence on whether smokers are more likely than non-smokers to experience negative progression or adverse outcomes from COVID-19 (3–5). Other unanswered questions are whether smokers are more worried than non-smokers about contracting or becoming ill from COVID-19, how smoking relates to adherence to protective behaviours, whether the amount people are smoking is changing in the context of the pandemic, and how far any of these factors are moderated by socioeconomic position. Understanding these issues is important for evaluating clinical risk, developing clear public health messaging, and identifying targets for intervention.

Former or current smoking increases the risk of a number of respiratory viral (6,7) and bacterial (8,9) infections and is associated with worse outcomes for those infected. Cigarette smoke reduces respiratory immune defences (10) and behavioural aspects of smoking (e.g. repeated hand-to-mouth action) may contribute to increased transmission of infectious agents (11). However, the evidence on smoking and COVID-19 risk specifically is less clear. Several studies have indicated that smokers may be more likely than non-smokers to develop severe respiratory disease from COVID-19 (3,4), but this has not been observed consistently (5). A recent review of 28 observational studies concluded there was limited evidence that disease severity in those hospitalised for COVID-19 is greater in current/former smokers than never smokers, but there was insufficient evidence to draw conclusions on infection, hospitalisation, or mortality (4). The number of cases in the descriptive studies conducted to date has typically been very small and lack of adjustment for relevant confounders means it is not possible to disentangle the effect of smoking (12). For example, smoking is known to increase the risk of chronic health conditions associated with poorer COVID-19 outcomes (e.g. hypertension, diabetes) (13–15).

Further complicating the picture are data from some countries, which suggest smoking prevalence is disproportionately low among hospitalised patients with COVID-19 (4,16,17). This is at odds with the higher rates of male versus female COVID-19 patients despite higher prevalence of smoking among men than women (13,18). It is not clear how far the underrepresentation of smokers among COVID-19 inpatients reflects underreporting of smoking (or conflation of former smoking with never smoking) in acute clinical settings, reverse causation (if smokers have stopped following the onset of COVID-19 symptoms), or self-selection (if smokers are less likely to present to hospital owing to lack of access to healthcare or more likely to die in the community from sudden complications) as opposed to a genuine protective effect of smoking (4,19). Population-level data collected outside of hospital settings are required.

How the pandemic is affecting smoking behaviour has implications for the provision and targeting of cessation support. On one hand, concerns about respiratory health may prompt some smokers to cut down or attempt to quit completely in order to reduce their risk of complications from COVID-19. It is not known whether smokers are more worried about developing COVID-19 or becoming seriously ill from the disease, which could affect their intention to quit (20). On the other hand, people may be smoking more than usual in an attempt to cope with higher than usual levels of stress (21–23) or to relieve boredom (23,24). Public health messaging may also influence smoking behaviour. Several organisations have warned of increased risk to smokers (25–27). Public Health England has advised smokers to quit to reduce their risk (27) and campaigns on social media are encouraging smokers to ‘#quit4covid’. However, these efforts may be undermined by headlines heralding a potential protective effect of smoking and nicotine (e.g., 28) based on reports of disproportionate hospitalisation rates (16,17), which has led to governments having to restrict the sale of nicotine replacement therapy to avoid panic buying (29).

In understanding associations between smoking and COVID-19, it is also important to consider the influence of socioeconomic position. Smoking is a socially patterned behaviour; the substantially higher prevalence of smoking in groups with greater socioeconomic disadvantage is a key driver of health inequalities (2,30). Despite continuing rhetoric that “we are all in this together”, the COVID-19 pandemic does not affect everyone equally (31). Front-line ‘key workers’ – comprising disproportionately lower paid roles, including healthcare, food supply chain, refuse collection, and public transport – are unable to stay at home or effectively practise social distancing and thus face greater risk of exposure to the virus. Others on low incomes are having to choose between adhering to protective behaviours (e.g. stay at home advice) and going out to work in order to afford basic outgoings. For disadvantaged smokers, greater exposure to stressors during the pandemic (e.g. financial strain, poor working and living conditions) may lead to compensatory increases in smoking for stress relief or make cutting down/quitting more difficult or a lower priority. Those who are motivated to quit are likely less able to access support from Stop Smoking Services, primary care or vape shops (which are closed). There may also be a social gradient associated with finding alternative support, especially digital. Understanding how socioeconomic position affects associations between smoking and COVID-19 relevant outcomes is essential for developing and targeting appropriate advice for smokers and evaluating the potential impact of interventions on health inequalities.

To summarise, there is a need for robust, population-based evidence on the association of smoking with COVID-19, taking into account the influence of inequalities and adjusting for potential confounding variables. Using cross-sectional data from a large study of adults in the UK, we aimed to address the following research questions:

1. Among adults in the UK, is smoking status associated with diagnosed or suspected COVID-19, after adjustment for sociodemographic characteristics, key worker status, and comorbid health conditions?
2. Among adults in the UK, is smoking status associated with worry or significant stress about contracting or becoming seriously ill with COVID-19, after adjustment for sociodemographic characteristics, key worker status, comorbid health conditions, and anxiety disorders?
3. Among adults in the UK, is smoking status associated with adherence to COVID-19 protective behaviours, after adjustment for sociodemographic characteristics, key worker status, and comorbid health conditions?
4. Among smokers, what proportions report smoking more than usual, about the same as usual, and less than usual over the past week?
5. Are recent changes in smoking associated with heaviness of smoking, sociodemographic characteristics, comorbid health conditions, confirmed or suspected COVID-19, or stress about becoming seriously ill with COVID-19?
6. Do the above associations differ by post-16 education as an indicator of socioeconomic position?

## Method

### Design

We used cross-sectional data from the UCL COVID-19 Social Study’s baseline survey. The COVID-19 Social Study is a longitudinal panel survey of adults (≥18 years) in the UK designed to provide insights into psychological and social experiences during the outbreak of SARS-CoV-2. The study sampling does not aim to be representative of the UK population, but is intended to have good representation across major sociodemographic groups. Thus, the sample has been recruited through a variety of channels including through the media, through targeted advertising by online advertising companies offering *pro-bono* support to ensure representation across major socio-demographic groups, and through partnerships with organisations representing vulnerable groups, enabling meaningful subgroup analysis. Participants complete a baseline survey, which assesses a range of information including demographics, health, smoking, confirmed and suspected COVID-19, and behaviours and attitudes relating to COVID-19, and are followed up weekly.

Data collection began on 21 March 2020. For this analysis, we aggregated data collected daily through 20 April 2020 (the most recent data available at the time of analysis).

### Patient and public involvement

The research questions in the UCL COVID-19 Social Study built on patient and public involvement as part of the UKRI MARCH Mental Health Research Network, which focuses on social, cultural and community engagement, and mental health. This highlighted priority research questions and measures for this study. Patients and the public were additionally involved in the recruitment of participants to the study and are actively involved in plans for the dissemination of findings from the study.

### Measures

#### Smoking status

Smoking status was assessed with the question: “Do you smoke?” with the response options: (a) nonsmoker, (b) ex-smoker, (c) current light smoker (9 or less a day), (d) current moderate smoker (10-19 a day), (e) current heavy smoker (20+ a day). For our primary analyses, participants answering (c), (d), or (e) were combined as ‘current smokers’.

#### Sociodemographic information

Highest level of education was used as a measure of socioeconomic position. This was analysed as a dichotomous variable: post-16 qualifications (completed post-16 vocational course, A-levels or equivalent [at school until age 18], undergraduate degree or professional qualification, postgraduate degree) vs. no post-16 qualifications (no qualifications, completed GCSE/CSE/O-levels or equivalent [at school until age 16]). We selected education over other markers of socioeconomic position (e.g. income or employment) because it provides a more reliable indication of socioeconomic position prior to COVID-19 (because it is not affected by recent job loss or furlough) and previous studies have shown level of education to be robustly associated with smoking status (32,33) and health outcomes (34).

Other sociodemographic variables included age (18-29, 30-39, 40-49, 50-59, 60-69, ≥70 years), sex, and ethnicity (white vs. other). These were included as covariates in our analyses given evidence that COVID-19 mortality rates are disproportionately high among older, male, and black and minority ethnic people (13,18,35). We also included information on key worker status, as this may be associated with increased exposure to COVID-19 (details of this measure are provided in Supplementary File 1).

#### Health conditions

The presence of smoking-associated health conditions was assessed with the question: “Do you have any of the following medical conditions?” Those who selected ‘high blood pressure’, ‘diabetes’, ‘heart disease’, ‘lung disease (e.g. asthma or COPD)’, or ‘cancer’ were coded 1 and those who selected none of these were coded 0. This variable was used as a covariate across the analyses, because people with chronic conditions may be more likely to experience severe symptoms (13–15).

The presence of anxiety disorders was assessed with the same question, with those who selected ‘clinically-diagnosed anxiety’ coded 1 and those who did not select this response coded 0. This variable was used as a covariate in analyses that include suspected COVID-19 and worry about COVID-19.

#### Confirmed and suspected COVID-19

Participants were asked: “Have you had COVID-19 (coronavirus)?” with response options (a) yes diagnosed and recovered, (b) yes diagnosed and still ill, (c) not formally diagnosed but suspected, (d) no. Confirmed COVID-19 was coded 1 for those who responded (a) or (b) and 0 for those who responded (c) or (d). Confirmed or suspected COVID-19 was coded 1 for those who responded (a), (b), or (c) and 0 for those who responded (d).

#### Worry about COVID-19

We assessed worry about COVID-19 with two questions which asked: “Over the past week, have any of the following been worrying you at all, even if only in a minor way?” and “Have any of these things been causing you significant stress? (e.g. they have been constantly on your mind or have been keeping you awake at night)”. Response options included ‘catching COVID-19’ and ‘becoming seriously ill from COVID-19’ among a range of other topics including social relationships, work, money, and getting food. We analysed four variables: (i) worry about catching COVID-19, (ii) significant stress about catching COVID-19, (iii) worry about becoming seriously ill from COVID-19 and (iv) significant stress about becoming seriously ill from COVID-19. For each variable, those who reported worry/stress about the relevant outcome were coded 1, else they were coded 0.

#### Adherence to COVID-19 protective behaviours

We used two items to assess adherence to COVID-19 protective behaviours. The primary measure was based on responses to the question: “Are you following the recommendation from authorities to prevent spread of COVID-19?” with responses on a scale from 1 (not at all) to 7 (very much so). Responses of 5 and above were coded 1 (indicating general adherence) and responses of 4 and below were coded 0.

As a secondary measure of (non)adherence, we examined the proportion who responded ‘I am living my life as normal’ (coded 1) in response to the question: “What is your current isolation status?”. Those who reported cutting down on usual activities, staying at home, and/or self-isolating were coded 0.

#### Recent changes in smoking

Past-week changes in smoking were assessed with the question: “Over the past week have you smoked more than usual?” with response options (a) less than usual, (b) about the same, (c) more than usual, (d) I don’t smoke. For analysis of smoking less than usual, smokers who responded (a) were coded 1 and those who responded (b) or (c) were coded 0. For analysis of smoking more than usual, smokers who responded (c) were coded 1 and those who responded (a) or (b) were coded 0. Those who responded (d) (i.e. never smokers and ex-smokers) were excluded from analysis of changes in smoking.

### Statistical analysis

The analysis plan was pre-registered on Open Science Framework (https://osf.io/pcs49/). Analyses were conducted on complete cases using SPSS v.24. To account for the non-random nature of the sample, all data were weighted to the proportions of sex, age, ethnicity, education, and country of living obtained from the Office for National Statistics (36).

We used logistic regression to examine associations between smoking status (never smoker [referent], ex-smoker, current smoker) and confirmed and suspected COVID-19, worry about COVID-19, and adherence to protective behaviours. For each outcome, we report the unadjusted association and adjusted models with sequential adjustment for relevant covariates (see Supplementary File 1 or table footnotes for full details of each model tested). Among current smokers, we used logistic regression to analyse unadjusted and multivariable associations of sociodemographics, comorbid health conditions, confirmed and suspected COVID-19, significant stress about becoming seriously ill from COVID-19, and survey date with recent changes in smoking.

## Results

A total of 55,481 participants responded to the survey between 21 March and 20 April 2020, of whom 53,221 (95.9%; weighted *n*=53,002) provided complete data on the variables included in the present analyses. Of the analysed sample, 13,602 (25.7%) were ex-smokers and 8,057 (15.2%) were current smokers. Sample characteristics in relation to smoking status are shown in Table 1. Relative to never smokers, current and ex-smokers were more likely to be middle-aged (current) or older (ex-smoker), male, white, have no post-16 qualifications, and have at least one chronic health condition. Current smokers were more likely than never smokers to be a key worker, and ex-smokers less likely. Current smokers were more likely than never smokers and ex-smokers to have a diagnosed anxiety disorder.

**Table 1.** Sample characteristics

|  | Whole sample | Never smokers | Ex-smokers | Current smokers | *p*<sup>1</sup> |
| --- | --- | --- | --- | --- | --- |
| *N* | 53002 | 31344 | 13602 | 8057 | - |
| Age in years, % (*n*) |  |  |  |  |  |
| 18-29 | 19.5 (10333) | 23.3 (7298) | 8.7 (1177) | 23.1 (1858) | <0.001 |
| 30-39 | 15.0 (7942) | 14.8 (4634) | 13.3 (1806) | 18.6 (1502) | - |
| 40-49 | 17.7 (9372) | 15.7 (4934) | 19.5 (2647) | 22.2 (1791) | - |
| 50-59 | 17.5 (9297) | 16.7 (5228) | 18.5 (2520) | 19.2 (1549) | - |
| 60-69 | 19.1 (10100) | 18.4 (5764) | 24.5 (3326) | 12.5 (1010) | - |
| ≥70 | 11.2 (5959) | 11.1 (3486) | 15.6 (2126) | 4.3 (347) | - |
| Female sex, % (*n*) | 50.6 (26825) | 52.4 (16432) | 45.9 (6245) | 51.5 (4148) | <0.001 |
| White ethnicity, % (*n*) | 87.2 (46219) | 85.5 (26789) | 91.0 (12378) | 87.5 (7052) | <0.001 |
| No post-16 qualifications, % (*n*) | 32.7 (17324) | 26.4 (8286) | 38.3 (5205) | 47.6 (3833) | <0.001 |
| Key worker, % (*n*) | 22.6 (11956) | 22.6 (7080) | 20.9 (2840) | 25.3 (2036) | <0.001 |
| ≥1 health condition<sup>2</sup>, % (*n*) | 32.2 (17093) | 28.8 (9029) | 40.9 (5557) | 31.1 (2507) | <0.001 |
| Diagnosed anxiety disorder, % (*n*) | 13.0 (6872) | 11.1 (3474) | 12.7 (1726) | 20.8 (1672) | <0.001 |
| Heavy smokers (≥10 CPD), % (*n*) | - | - | - | 50.3 (4056) | - |

Note: All data are weighted to match the UK adult population on sex, age, ethnicity, education, and country of living. CPD, cigarettes per day.
<sup>1</sup> *p* value for the association between each variable and smoking status.
<sup>2</sup> High blood pressure, diabetes, heart disease, lung disease (e.g. asthma or COPD), or cancer.

### Confirmed and suspected COVID-19

Of the sample, 0.3% [95% confidence interval (CI) 0.3-0.3%] reported having (had) a confirmed case of COVID-19 and a further 10.3% [95%CI 10.0-10.5%] reported experiencing symptoms of COVID-19. Compared with never smokers, prevalence of confirmed COVID-19 was higher among current smokers but not ex-smokers, and the associations were similar after adjustment. Associations between smoking status and confirmed and suspected COVID-19 differed significantly by level of education. Odds of confirmed COVID-19 were 3.5 times higher among current smokers with no post-16 qualifications than never smokers after adjustment for covariates, but did not differ significantly by smoking status among participants with post-16 qualifications (Table 2).

**Table 2.** Association of smoking status with confirmed and suspected COVID-19

|  | %<br>[95% CI] | Model 1 |  | Model 2 |  | Model 3 |  | Model 4 |  | Interaction with post-16 qualifications<sup>1</sup> |  |
| --- | --- | --- | --- | --- | --- | --- | --- | --- | --- | --- | --- |
|  |  | OR<br>[95% CI] | *p* | OR<sub>adj</sub><br>[95% CI] | *p* | OR<sub>adj</sub><br>[95% CI] | *p* | OR<sub>adj</sub><br>[95% CI] | *p* | OR<sub>adj</sub><br>[95% CI] | *p* |
| Confirmed COVID-19 |  |  |  |  |  |  |  |  |  |  |  |
| Whole sample |  |  |  |  |  |  |  |  |  |  |  |
| Never smoker | 0.3<br>[0.2-0.3] | - | - | - | - | - | - | - | - | - | - |
| Ex-smoker | 0.2<br>[0.2-0.3] | 0.73<br>[0.47-1.14] | 0.163 | 0.89<br>[0.57-1.40] | 0.615 | 0.85<br>[0.54-1.33] | 0.465 | - | - | 0.26<br>[0.09-0.79] | 0.017 |
| Current smoker | 0.6<br>[0.4-0.8] | 2.14<br>[1.49-3.08] | <0.001 | 1.87<br>[1.28-2.73] | 0.001 | 1.79<br>[1.22-2.62] | 0.003 | - | - | 3.74<br>[1.51-9.27] | 0.004 |
| No post-16 qualifications |  |  |  |  |  |  |  |  |  |  |  |
| Never smoker | 0.3<br>[0.2-0.5] | - | - | - | - | - | - | - | - | - | - |
| Ex-smoker | 0.1<br>[0.0-0.2] | 0.28<br>[0.10-0.76] | 0.012 | 0.49<br>[0.18-1.37] | 0.175 | 0.49<br>[0.18-1.37] | 0.172 | - | - | - | - |
| Current smoker | 1.0<br>[0.7-1.4] | 3.16<br>[1.91-5.21] | <0.001 | 3.64<br>[2.11-6.28] | <0.001 | 3.53<br>[2.04-6.10] | <0.001 | - | - | - | - |
| Post-16 qualifications |  |  |  |  |  |  |  |  |  |  |  |
| Never smoker | 0.2<br>[0.2-0.3] | - | - | - | - | - | - | - | - | - | - |
| Ex-smoker | 0.3<br>[0.2-0.4] | 1.05<br>[0.64-1.72] | 0.850 | 1.14<br>[0.69-1.88] | 0.618 | 1.04<br>[0.63-1.72] | 0.888 | - | - | - | - |
| Current smoker | 0.2<br>[0.1-0.4] | 0.76<br>[0.36-1.59] | 0.461 | 0.69<br>[0.33-1.47] | 0.337 | 0.67<br>[0.32-1.43] | 0.302 | - | - | - | - |

**Table 2.** (continued)
|  |  | Model 1 |  | Model 2 |  | Model 3 |  | Model 4 |  | Interaction with post 16 qualifications<sup>1</sup> |  |
| --- | --- | --- | --- | --- | --- | --- | --- | --- | --- | --- | --- |
|  | %<br>[95% CI] | OR<br>[95% CI] | *p* | OR<sub>adj</sub><br>[95% CI] | *p* | OR<sub>adj</sub><br>[95% CI] | *p* | OR<sub>adj</sub><br>[95% CI] | *p* | OR<sub>adj</sub><br>[95% CI] | *p* |
| Confirmed and suspected COVID-19 |  |  |  |  |  |  |  |  |  |  |  |
| Whole sample |  |  |  |  |  |  |  |  |  |  |  |
| Never smoker | 10.2<br>[9.9-10.6] | - | - | - | - | - | - | - | - | - | - |
| Ex-smoker | 10.9<br>[10.4-11.5] | 1.07<br>[1.01-1.15] | 0.033 | 1.22<br>[1.14-1.31] | <0.001 | 1.22<br>[1.14-1.31] | <0.001 | 1.21<br>[1.13-1.29] | <0.001 | 0.26<br>[0.09-0.79] | 0.018 |
| Current smoker | 11.2<br>[10.6-11.9] | 1.11<br>[1.03-1.20] | 0.009 | 1.05<br>[0.97-1.14] | 0.199 | 1.06<br>[0.97-1.14] | 0.197 | 1.02<br>[0.94-1.11] | 0.572 | 3.77<br>[1.52-9.35] | 0.004 |
| No post-16 qualifications |  |  |  |  |  |  |  |  |  |  |  |
| Never smoker | 8.0<br>[7.4-8.6] | - | - | - | - | - | - | - | - | - | - |
| Ex-smoker | 7.9<br>[7.2-8.6] | 0.99<br>[0.87-1.12] | 0.850 | 1.03<br>[0.91-1.18] | 0.625 | 1.03<br>[0.91-1.18] | 0.620 | 1.02<br>[0.90-1.17] | 0.747 | - | - |
| Current smoker | 10.8<br>[9.9-11.9] | 1.40<br>[1.23-1.60] | <0.001 | 1.16<br>[1.01-1.32] | 0.032 | 1.16<br>[1.01-1.32] | 0.032 | 1.13<br>[0.98-1.29] | 0.083 | - | - |
| Post-16 qualifications |  |  |  |  |  |  |  |  |  |  |  |
| Never smoker | 11.1<br>[10.7-11.5] | - | - | - | - | - | - | - | - | - | - |
| Ex-smoker | 12.8<br>[12.1-13.5] | 1.18<br>[1.09-1.27] | <0.001 | 1.31<br>[1.21-1.42] | <0.001 | 1.31<br>[1.21-1.42] | <0.001 | 1.30<br>[1.20-1.40] | <0.001 | - | - |
| Current smoker | 11.6<br>[10.7-12.6] | 1.06<br>[0.95-1.17] | 0.302 | 0.96<br>[0.87-1.07] | 0.474 | 0.96<br>[0.87-1.07] | 0.477 | 0.94<br>[0.84-1.04] | 0.216 | - | - |

Note: All data are weighted to match the UK adult population on sex, age, ethnicity, education, and country of living. CI, confidence interval. OR, odds ratio. OR<sub>adj</sub>, adjusted odds ratio. Shaded rows are results of analyses stratified by post-16 qualifications.
Model 1 is unadjusted. Model 2 is adjusted for age, sex, ethnicity, post-16 qualifications, key worker status, and survey date. Model 3 is adjusted for age, sex, ethnicity, post-16 qualifications, key worker status, survey date, and comorbid health conditions. Model 4 is adjusted for age, sex, ethnicity, post-16 qualifications, key worker status, survey date, comorbid health conditions, and diagnosed anxiety disorders.
<sup>1</sup> Interaction between smoking status and post-16 qualifications added to fully adjusted model with the exclusion of key worker status.

After including suspected cases, prevalence was higher among current smokers and ex-smokers than never smokers, but remained higher only among ex-smokers after adjustment. Odds of confirmed and suspected COVID-19 were also significantly higher among current smokers with no post-16 qualifications after adjustment for sociodemographics, key worker status, and comorbid health conditions (OR=1.16) but this association was attenuated when diagnosed anxiety disorders were controlled for (OR=1.13; Table 2). Odds of confirmed and suspected COVID-19 were significantly higher among ex-smokers with post-16 qualifications than never smokers after full adjustment for covariates (OR=1.30; Table 2).

### Worry about COVID-19

Just under half of participants reported being worried about catching (45.1% [95%CI 44.7-45.6%]) or becoming seriously ill from COVID-19 (46.0% [95%CI 45.6-46.4%]), and one in five reported significant stress about these possibilities (19.1% [95%CI 18.7-19.4%] and 22.9% [95%CI 22.5-23.2%], respectively). Current and ex-smokers were significantly more likely than never smokers to report COVID-19 causing them worry or stress (Table 3). The association between smoking and worry about catching COVID-19 was stronger for smokers with no post-16 qualifications than those with post-16 qualifications, but there was no interaction with level of education for associations between smoking status and worry about becoming seriously ill from COVID-19 or significant stress about catching or becoming seriously ill from COVID-19 (Table 3).

**Table 3.** Association of smoking status with worry about COVID-19

|  | %<br>[95% CI] | Model 1 |  | Model 2 |  | Model 3 |  | Interaction with post-16 qualifications<sup>1</sup> |  |
| --- | --- | --- | --- | --- | --- | --- | --- | --- | --- |
|  |  | OR<br>[95% CI] | *p* | OR<sub>adj</sub><br>[95% CI] | *p* | OR<sub>adj</sub><br>[95% CI] | *p* | OR<sub>adj</sub><br>[95% CI] | *p* |
| Worry about catching COVID-19 |  |  |  |  |  |  |  |  |  |
| Whole sample |  |  |  |  |  |  |  |  |  |
| Never smoker | 43.4<br>[42.8-43.9] | - | - | - | - | - | - | - | - |
| Ex-smoker | 46.5<br>[45.6-47.3] | 1.13<br>[1.09-1.18] | <0.001 | 1.14<br>[1.09-1.19] | <0.001 | 1.10<br>[1.06-1.15] | <0.001 | 1.22<br>[1.12-1.33] | <0.001 |
| Current smoker | 49.8<br>[48.7-50.9] | 1.30<br>[1.23-1.36] | <0.001 | 1.26<br>[1.19-1.32] | <0.001 | 1.20<br>[1.14-1.26] | <0.001 | 1.10<br>[1.00-1.22] | 0.063 |
| No post-16 qualifications |  |  |  |  |  |  |  |  |  |
| Never smoker | 43.0<br>[41.9-44.1] | - | - | - | - | - | - | - | - |
| Ex-smoker | 49.0<br>[47.7-50.4] | 1.28<br>[1.19-1.37] | <0.001 | 1.27<br>[1.18-1.36] | <0.001 | 1.24<br>[1.15-1.33] | <0.001 | - | - |
| Current smoker | 51.3<br>[49.1-52.2] | 1.40<br>[1.29-1.51] | <0.001 | 1.31<br>[1.21-1.42] | <0.001 | 1.26<br>[1.16-1.36] | <0.001 | - | - |
| Post-16 qualifications |  |  |  |  |  |  |  |  |  |
| Never smoker | 43.5<br>[42.9-44.2] | - | - | - | - | - | - | - | - |
| Ex-smoker | 44.9<br>[43.8-45.9] | 1.06<br>[1.00-1.11] | 0.033 | 1.07<br>[1.02-1.13] | 0.006 | 1.04<br>[0.98-1.09] | 0.178 | - | - |
| Current smoker | 48.4<br>[46.9-49.9] | 1.22<br>[1.14-1.30] | <0.001 | 1.22<br>[1.14-1.30] | <0.001 | 1.17<br>[1.10-1.25] | <0.001 | - | - |

Table 3. (continued)
|  | %<br>[95% CI] | Model 1 |  | Model 2 |  | Model 3 |  | Interaction with post-16 qualifications<sup>1</sup> |  |
| --- | --- | --- | --- | --- | --- | --- | --- | --- | --- |
|  |  | OR<br>[95% CI] | *p* | OR<sub>adj</sub><br>[95% CI] | *p* | OR<sub>adj</sub><br>[95% CI] | *p* | OR<sub>adj</sub><br>[95% CI] | *p* |
| Significant stress about catching COVID-19 |  |  |  |  |  |  |  |  |  |
| Whole sample |  |  |  |  |  |  |  |  |  |
| Never smoker | 16.8<br>[16.4-17.3] | - | - | - | - | - | - | - | - |
| Ex-smoker | 19.9<br>[19.2-20.6] | 1.23<br>[1.17-1.29] | <0.001 | 1.22<br>[1.16-1.29] | <0.001 | 1.15<br>[1.09-1.22] | <0.001 | 1.07<br>[0.96-1.20] | 0.211 |
| Current smoker | 26.3<br>[25.4-27.3] | 1.77<br>[1.67-1.87] | <0.001 | 1.56<br>[1.47-1.66] | <0.001 | 1.43<br>[1.35-1.52] | <0.001 | 1.00<br>[0.89-1.13] | 0.998 |
| Worry about becoming seriously ill from COVID-19 |  |  |  |  |  |  |  |  |  |
| Whole sample |  |  |  |  |  |  |  |  |  |
| Never smoker | 44.0<br>[43.4-44.5] | - | - | - | - | - | - | - | - |
| Ex-smoker | 48.8<br>[48.0-49.6] | 1.21<br>[1.17-1.26] | <0.001 | 1.25<br>[1.20-1.30] | <0.001 | 1.19<br>[1.14-1.24] | <0.001 | 1.04<br>[0.95-1.14] | 0.395 |
| Current smoker | 49.2<br>[48.1-50.3] | 1.23<br>[1.18-1.30] | <0.001 | 1.23<br>[1.17-1.29] | <0.001 | 1.15<br>[1.09-1.21] | <0.001 | 1.08<br>[0.97-1.19] | 0.168 |
| Significant stress about becoming seriously ill from COVID-19 |  |  |  |  |  |  |  |  |  |
| Whole sample |  |  |  |  |  |  |  |  |  |
| Never smoker | 20.4<br>[20.0-20.9] | - | - | - | - | - | - | - | - |
| Ex-smoker | 24.7<br>[24.0-25.4] | 1.28<br>[1.22-1.34] | <0.001 | 1.31<br>[1.24-1.37] | <0.001 | 1.22<br>[1.16-1.28] | <0.001 | 1.01<br>[0.91-1.12] | 0.880 |
| Current smoker | 29.4<br>[28.4-30.4] | 1.62<br>[1.53-1.71] | <0.001 | 1.49<br>[1.40-1.57] | <0.001 | 1.34<br>[1.27-1.43] | <0.001 | 1.09<br>[0.97-1.23] | 0.156 |

Note: All data are weighted to match the UK adult population on sex, age, ethnicity, education, and country of living. CI, confidence interval. OR, odds ratio. OR<sub>adj</sub>, adjusted odds ratio. Shaded rows are results of analyses stratified by post-16 qualifications.
Model 1 is unadjusted. Model 2 is adjusted for age, sex, ethnicity, post-16 qualifications, key worker status, and survey date. Model 3 is adjusted for age, sex, ethnicity, post-16 qualifications, key worker status, survey date, comorbid health conditions, and diagnosed anxiety disorders.
<sup>1</sup> Interaction between smoking status and post-16 qualifications added to fully adjusted model with the exclusion of key worker status.

### Adherence to COVID-19 protective behaviours

Adherence to COVID-19 protective behaviours was very high: 96.3% [95%CI 96.1-96.4%] reported general adherence to recommendations from authorities to prevent the spread of COVID-19, and just 3.7% [95%CI 3.5-3.8%] reported living life as normal. Associations between smoking status and adherence to protective behaviours differed significantly by level of education. Current smokers had lower odds of reporting general adherence relative to never smokers irrespective of education, but the association was stronger among those with post-16 qualifications (Table 4). Current smokers with post-16 qualifications also had higher odds of reporting living life as normal (i.e. lower odds of adherence) than never smokers, but there was no difference between current smokers and never smokers with no post-16 qualifications (Table 4). Ex-smokers with no post-16 qualifications were less likely than never smokers and current smokers to report living life as normal (i.e. more likely to adhere to recommendations; Table 4).

**Table 4.**
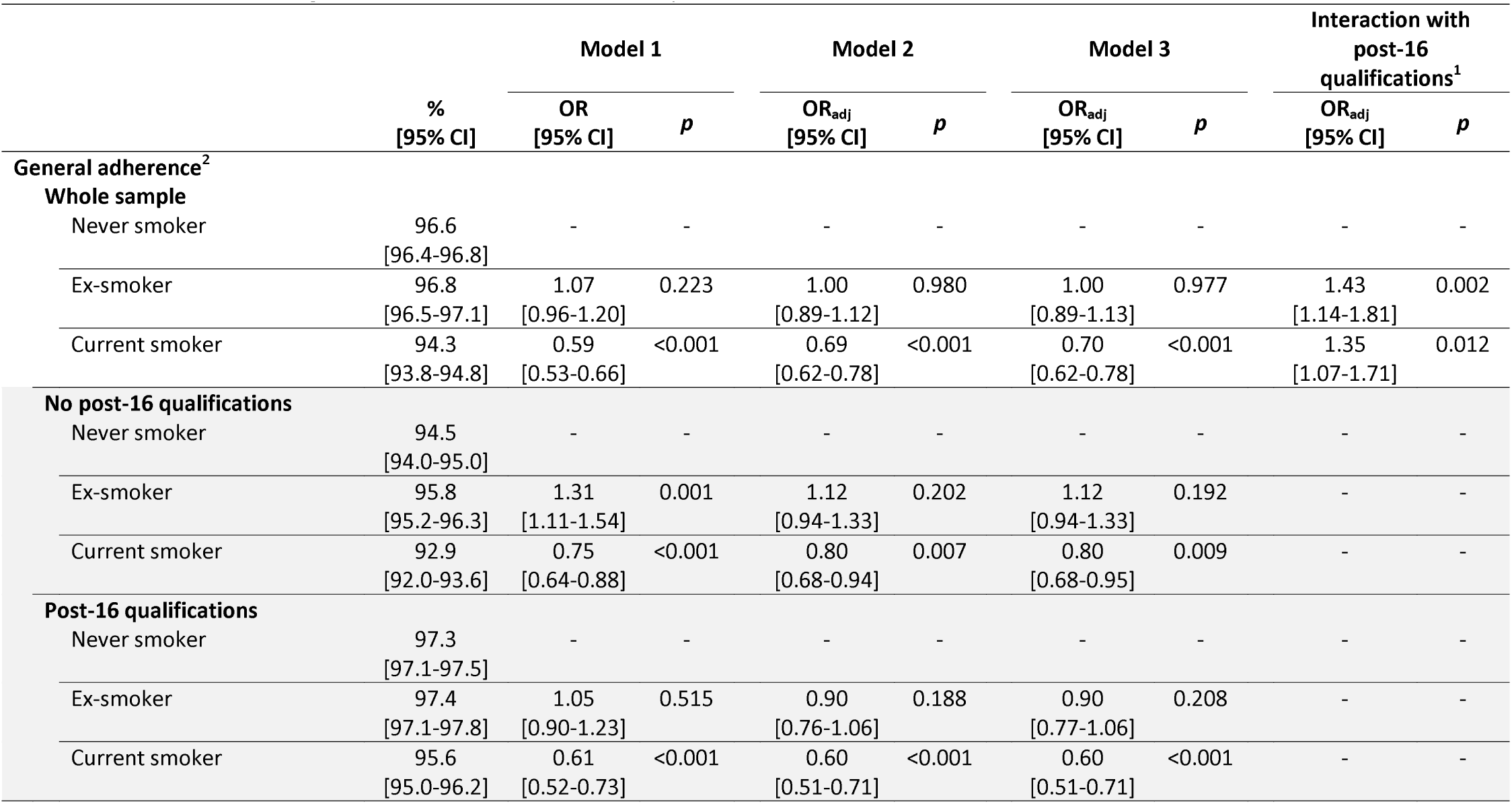
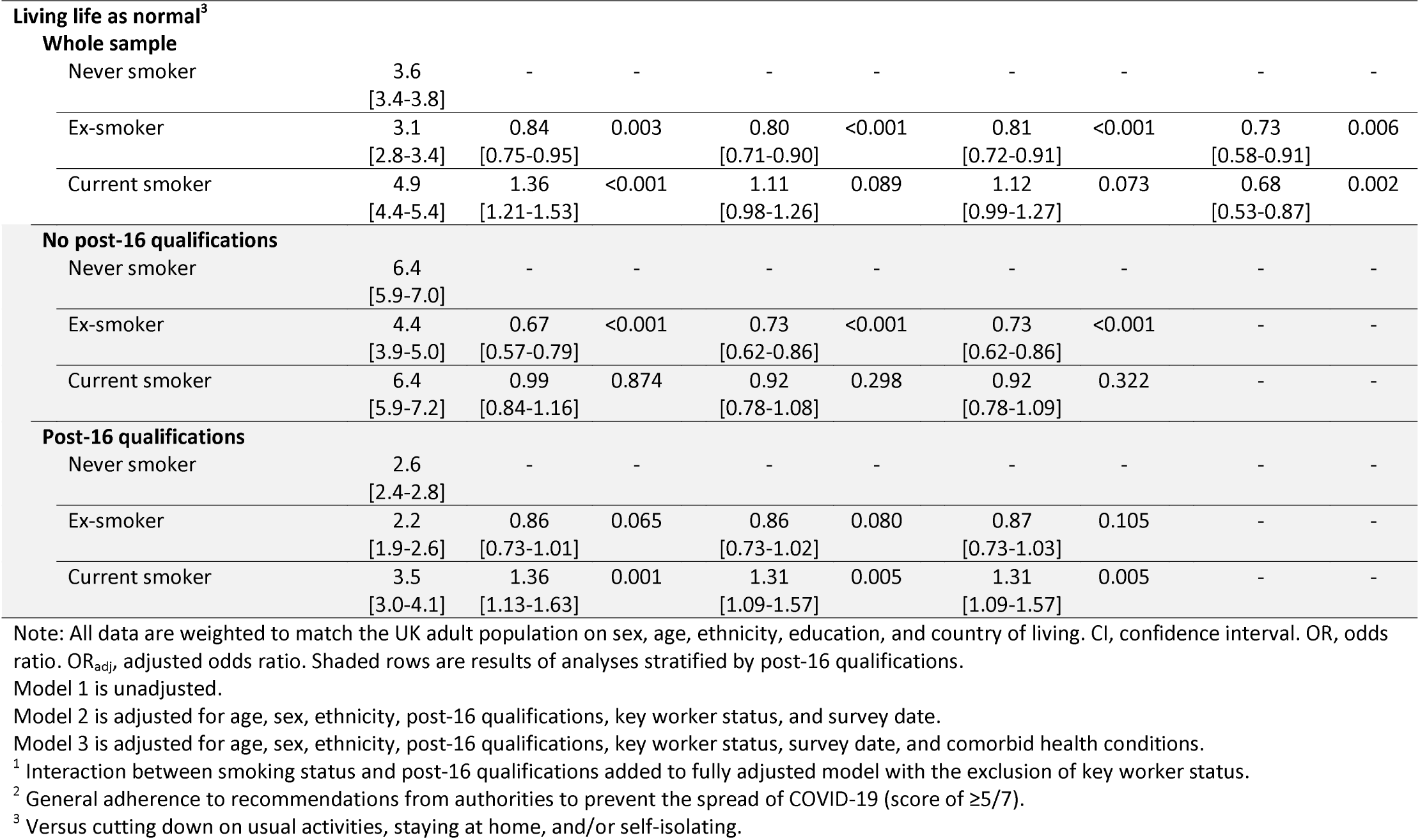
Association of smoking status with adherence to COVID-19 protective behaviours

### Recent changes in smoking

Among current smokers, 13.4% [95%CI 12.7-14.2%] reported smoking less than usual in the past week, 42.2% [95%CI 41.3-43.5%] reported smoking more than usual, and 43.9% [95%CI 43.0-45.1%] reported smoking about the same amount as usual. Smoking less than usual was independently associated with being a light smoker (<10 cigarettes per day), younger age, non-white ethnicity, having post-16 qualifications, and confirmed or suspected COVID-19 (Table 5). Among smokers with no post-16 qualifications, smoking less than usual was also associated with younger age (18-29 vs. ≥70 years), male sex, being a key worker, and absence of comorbid health conditions (Table 5). Among smokers with post-16 qualifications, smoking less than usual was also associated with not being a key worker, and experiencing significant stress about becoming seriously ill from COVID-19 (Table 5).

**Table 5.**
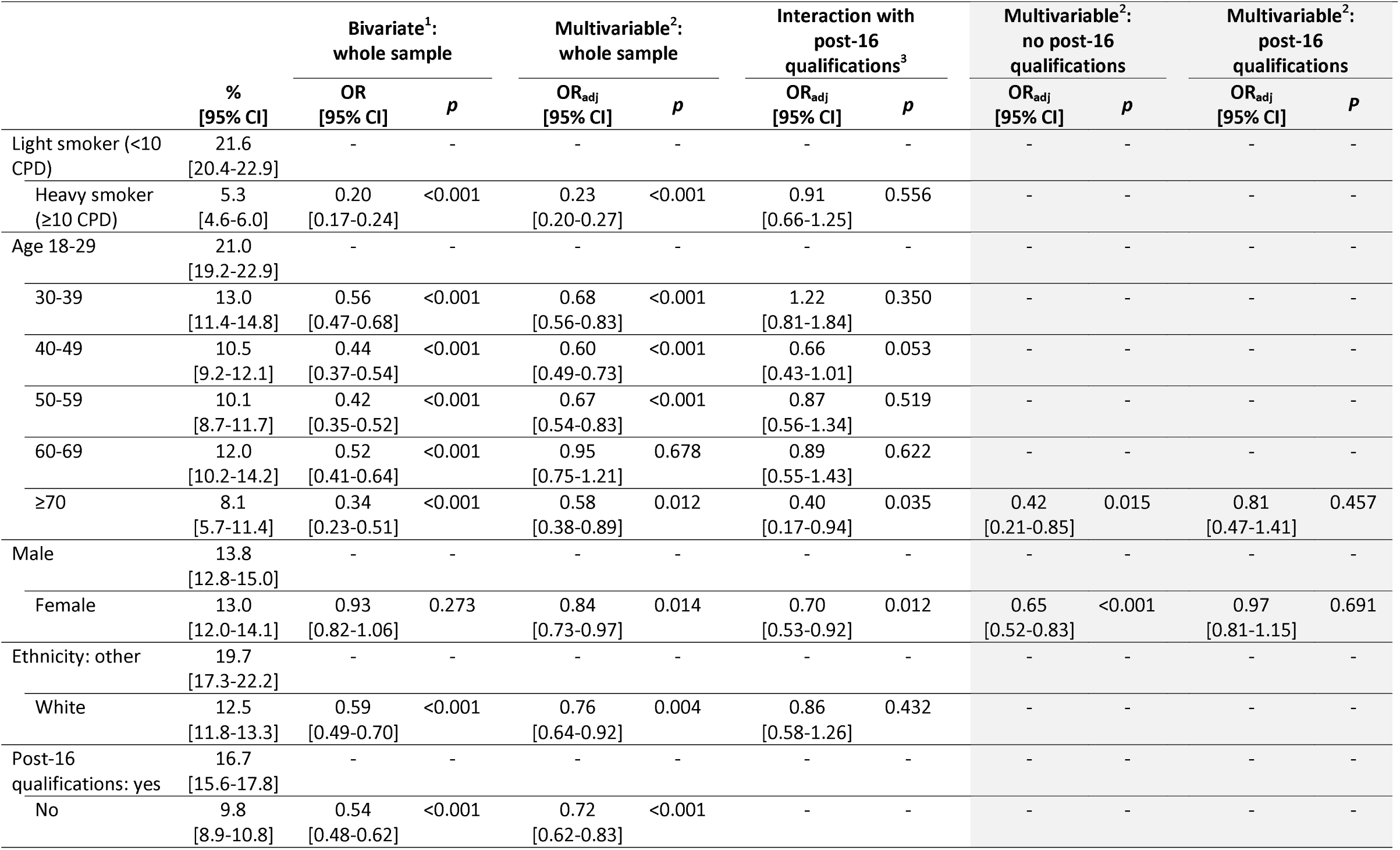
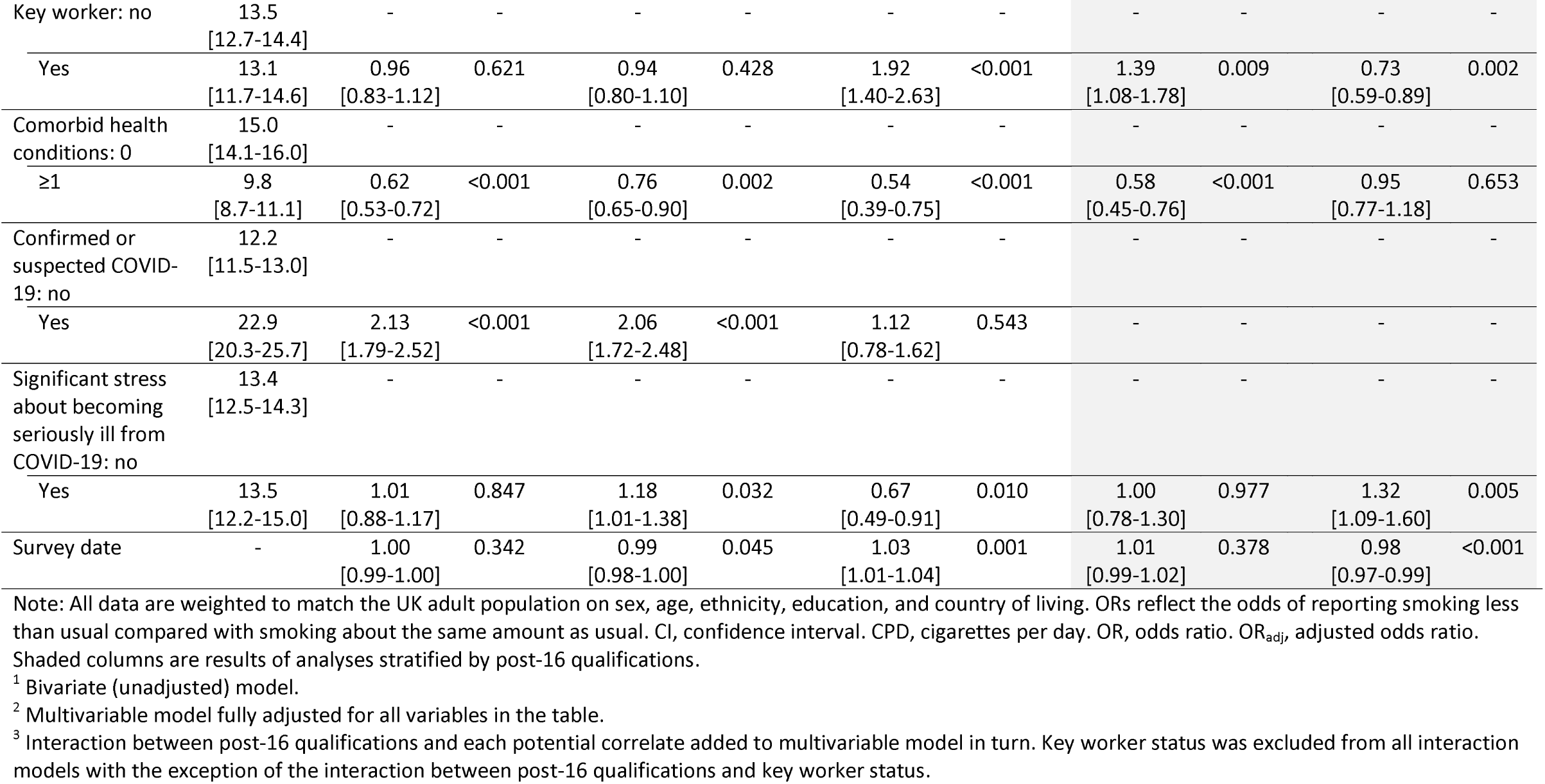
Correlates of smoking less than usual in the past week among current smokers

Smoking more than usual was independently associated with being a heavy smoker (≥10 cigarettes per day), younger, female, and experiencing significant stress about becoming seriously ill from COVID-19 (Table 6). Among smokers with no post-16 qualifications, smoking more than usual was also associated with not being a key worker, having comorbid health conditions, and having confirmed or suspected COVID-19 (Table 6). Among smokers with post-16 qualifications, smoking more than usual was also associated with not having confirmed or suspected COVID-19 (Table 6). Associations of smoking more than usual with female sex and experience significant stress about becoming seriously ill from COVID-19 were stronger for smokers who had no post-16 qualifications than those who did (Table 6).

**Table 6.** Correlates of smoking more than usual in the past week among current smokers

|  | %<br>[95% CI] | Bivariate<sup>1</sup>:<br>whole sample |  | Multivariable<sup>2</sup>:<br>whole sample |  | Interaction with<br>post-16<br>qualifications<sup>3</sup> |  | Multivariable<sup>2</sup>:<br>no post-16<br>qualifications |  | Multivariable<sup>2</sup>:<br>post-16<br>qualifications |  |
| --- | --- | --- | --- | --- | --- | --- | --- | --- | --- | --- | --- |
|  |  | OR<br>[95% CI] | *p* | OR<sub>adj</sub><br>[95% CI] | *p* | OR<sub>adj</sub><br>[95% CI] | *p* | OR<sub>adj</sub><br>[95% CI] | *p* | OR<sub>adj</sub><br>[95% CI] | *p* |
| Light smoker (<10 CPD) | 35.5<br>[34.0-37.0] | - | - | - | - | - | - | - | - | - | - |
| Heavy smoker (≥10 CPD) | 49.2<br>[47.7-50.8] | 1.77<br>[1.62-1.93] | <0.001 | 2.07<br>[1.87-2.28] | <0.001 | 0.98<br>[0.81-1.19] | 0.836 | - | - | - | - |
| Age 18-29 | 49.2<br>[46.9-51.5] | - | - | - | - | - | - | - | - | - | - |
| 30-39 | 51.1<br>[48.6-53.6] | 1.08<br>[0.94-1.24] | 0.279 | 0.98<br>[0.85-1.13] | 0.801 | 0.83<br>[0.62-1.11] | 0.209 | - | - | - | - |
| 40-49 | 41.6<br>[39.4-43.9] | 0.74<br>[0.65-0.84] | <0.001 | 0.62<br>[0.54-0.71] | <0.001 | 0.82<br>[0.62-1.08] | 0.152 | - | - | - | - |
| 50-59 | 36.9<br>[34.5-39.3] | 0.60<br>[0.53-0.69] | <0.001 | 0.48<br>[0.42-0.56] | <0.001 | 1.04<br>[0.77-1.40] | 0.801 | - | - | - | - |
| 60-69 | 33.7<br>[30.9-36.7] | 0.53<br>[0.45-0.62] | <0.001 | 0.41<br>[0.34-0.48] | <0.001 | 0.86<br>[0.62-1.21] | 0.386 | - | - | - | - |
| ≥70 | 22.3<br>[18.2-26.9] | 0.30<br>[0.23-0.39] | <0.001 | 0.24<br>[0.18-0.33] | <0.001 | 0.73<br>[0.41-1.29] | 0.282 | - | - | - | - |
| Male | 37.5<br>[36.0-39.0] | - | - | - | - | - | - | - | - | - | - |
| Female | 47.0<br>[45.5-48.5] | 1.47<br>[1.35-1.61] | <0.001 | 1.33<br>[1.21-1.46] | <0.001 | 1.26<br>[1.04-1.52] | 0.016 | 1.47<br>[1.28-1.69] | <0.001 | 1.23<br>[1.08-1.40] | 0.002 |
| Ethnicity: other | 38.5<br>[35.6-41.6] | - | - | - | - | - | - | - | - | - | - |
| White | 42.9<br>[41.8-44.1] | 1.20<br>[1.05-1.38] | 0.009 | 1.09<br>[0.95-1.26] | 0.231 | 1.30<br>[0.97-1.74] | 0.085 | - | - | - | - |
| Post-16 qualifications: yes | 42.7<br>[41.2-44.2] | - | - | - | - | - | - | - | - | - | - |
| No | 42.0<br>[40.5-43.6] | 0.97<br>[0.89-1.06] | 0.529 | 0.91<br>[0.83-1.00] | 0.051 | - | - | - | - | - | - |

Table 6. (continued)
|  | %<br>[95% CI] | Bivariate<sup>1</sup>:<br>whole sample |  | Multivariable<sup>2</sup>:<br>whole sample |  | Interaction with<br>post-16<br>qualifications<sup>3</sup> |  | Multivariable<sup>2</sup>:<br>no post-16<br>qualifications |  | Multivariable<sup>2</sup>:<br>post-16<br>qualifications |  |
| --- | --- | --- | --- | --- | --- | --- | --- | --- | --- | --- | --- |
|  |  | OR<br>[95% CI] | *p* | OR<sub>adj</sub><br>[95% CI] | *p* | OR<sub>adj</sub><br>[95% CI] | *p* | OR<sub>adj</sub><br>[95% CI] | *p* | OR<sub>adj</sub><br>[95% CI] | *p* |
| Key worker: no | 43.2<br>[41.9-44.4] | - | - | - | - | - | - | - | - | - | - |
| Yes | 40.0<br>[37.9-42.2] | 0.88<br>[0.80-0.98] | 0.017 | 0.79<br>[0.71-0.88] | <0.001 | 0.63<br>[0.50-0.78] | <0.001 | 0.59<br>[0.50-0.70] | <0.001 | 0.98<br>[0.85-1.13] | 0.772 |
| Comorbid health conditions: 0 | 42.3<br>[41.0-43.6] | - | - | - | - | - | - | - | - | - | - |
| ≥1 | 42.6<br>[40.7-44.5] | 1.01<br>[0.92-1.11] | 0.810 | 1.18<br>[1.06-1.31] | 0.003 | 1.51<br>[1.23-1.86] | <0.001 | 1.42<br>[1.22-1.66] | <0.001 | 0.96<br>[0.82-1.12] | 0.601 |
| Confirmed or suspected COVID-19: no | 42.1<br>[41.0-43.3] | - | - | - | - | - | - | - | - | - | - |
| Yes | 44.4<br>[41.2-47.7] | 1.10<br>[0.96-1.26] | 0.189 | 0.94<br>[0.81-1.09] | 0.420 | 1.81<br>[1.35-2.43] | <0.001 | 1.30<br>[1.04-1.62] | 0.019 | 0.72<br>[0.59-0.88] | 0.001 |
| Significant stress about becoming seriously ill from COVID-19: no | 37.9<br>[36.7-39.2] | - | - | - | - | - | - | - | - | - | - |
| Yes | 53.2<br>[51.2-55.2] | 1.86<br>[1.69-2.05] | <0.001 | 1.84<br>[1.66-2.04] | <0.001 | 1.66<br>[1.35-2.03] | <0.001 | 2.35<br>[2.02-2.73] | <0.001 | 1.48<br>[1.28-1.71] | <0.001 |
| Survey date | - | 1.03<br>[1.02-1.03] | <0.001 | 1.03<br>[1.02-1.03] | <0.001 | 1.00<br>[0.99-1.01] | 0.524 | - | - | - | - |

Note: All data are weighted to match the UK adult population on sex, age, ethnicity, education, and country of living. ORs reflect the odds of reporting smoking more than usual compared with smoking about the same amount as usual. CI, confidence interval. CPD, cigarettes per day. OR, odds ratio. OR<sub>adj</sub>, adjusted odds ratio. Shaded columns are results of analyses stratified by post-16 qualifications.
<sup>1</sup> Bivariate (unadjusted) model.
<sup>2</sup> Multivariable model fully adjusted for all variables in the table.
<sup>3</sup> Interaction between post-16 qualifications and each potential correlate added to multivariable model in turn. Key worker status was excluded from all interaction models with the exception of the interaction between post-16 qualifications and key worker status.

## Discussion

In this large survey of adults living in the UK, current smoking was associated with 1.8 times higher odds of confirmed COVID-19 relative to never smoking, independent of age, sex, ethnicity, key worker status, and comorbid health conditions. This was driven by a substantially higher rate of confirmed COVID-19 among smokers with no post-16 qualifications, with no significant difference by smoking status observed in those who had post-16 qualifications. It should be noted that the low prevalence of confirmed cases in our sample (0.3%) resulted in wide confidence intervals. Nonetheless, our data provide no evidence to support a protective effect of smoking, in contrast with data from several countries documenting substantially lower smoking prevalence among hospitalised patients with COVID-19 than would be expected based on smoking prevalence in the population (4,16,17).

There was no significant difference between current smokers and never smokers when confirmed cases were combined with suspected cases of COVID-19, with higher raw prevalence among current smokers accounted for by sociodemographic characteristics, comorbid health conditions, and anxiety disorders. However, former smokers who had post-16 qualifications had 30% higher odds of confirmed or suspected COVID-19 even after full adjustment for covariates. It is possible that this association was driven by high rates of smoking cessation following the onset of COVID-19 symptoms (i.e. reverse causation) (4,19). In tentative support of this theory, our analyses showed that smokers with confirmed or suspected COVID-19 had twice the odds of reporting smoking less than usual in the past week than those without COVID-19. Data from a hospital in France also indicate that while prevalence of current smoking was lower in hospitalised COVID-19 patients than the general population, prevalence of former smoking was much higher and prevalence of never smoking similar (37).

It has been theorised that there may be a potential protective effect of smoking on COVID-19 outcomes, possibly via the interaction of nicotine with the renin-angiotensin system or the effects of nicotine on the immune system (16,38–40). However, the present results suggest the protection would need to be conferred independently of infection risk. Alternatively, there are other explanations for the low smoking rates among hospitalised patients. There are several possibilities, which are not mutually exclusive. Smokers quitting post-symptom onset – particularly among those with symptoms severe enough to warrant hospital treatment – may inflate the proportion of former smokers relative to current smokers. Recording smoking status is not likely to be high priority in acute clinical settings stretched to capacity in the midst of a pandemic, so prevalence of current smoking may be underestimated in hospital records or former smoking conflated with never smoking. Self-selection bias may also be present, whereby smokers are less likely to present to hospital because they lack funds to pay for medical care (2) or are more likely to suffer fatal complications in the community. As further population-based data become available, we will gain a clearer picture of true differences in infection rates between current smokers, former smokers, and never smokers. The Smoking Toolkit Study (41), which surveys a different representative sample of adults in England each month, has begun to collect data on COVID-19 infection which will allow more detailed assessment of associations between smoking and COVID-19 in the near future.

Beyond differences in the odds of COVID-19 infection, we also observed associations between smoking status and other COVID-19 relevant variables. Adherence to recommendations from authorities to prevent the spread of COVID-19 was generally high (96.3%), but current smokers had lower odds of reporting adherence than never smokers. This discrepancy was more pronounced among smokers with post-16 qualifications, although absolute rates of adherence were higher in current smokers with post-16 qualifications (95.6%) than never smokers with no post-16 qualifications (94.5%). Despite being less likely to adhere to protective behaviours, current and former smokers were significantly more likely to report worry or significant stress about catching or becoming seriously ill from COVID-19. This apparent contradiction between motivation and behaviour may reflect inequality in the opportunity or ability to adhere to guidelines. For some smokers with post-16 qualifications, the increased worry or stress may have translated to their increased efforts to cut down cigarette consumption, but for others with and without post-16 qualifications it was associated with smoking more, possibly in an effort to reduce or cope with stress (21–23,42). Further evidence on smoking potentially being used as a maladaptive coping mechanism by those with no post-16 qualifications is apparent in the data linking confirmed/suspected COVID-19 with changes in smoking: while COVID-19 cases had twice the odds of reporting reduced smoking relative to those without suspected COVID-19, those with no post-16 qualifications also had significantly increased odds of reporting smoking more than usual (an association not seen in smokers with post-16 qualifications). Among the current smokers we surveyed, a considerably higher proportion reported a recent increase in smoking (42.0%) than reported smoking less than usual (13.4%). Reduced smoking was more common among those with (16.7%) than without post-16 qualifications (9.8%). These findings emphasise the need for continued investment in tobacco control activity and cessation support during the pandemic, with focused effort to target disadvantaged smokers. Correcting the common misperception that smoking is effective in relieving stress could help to combat an exacerbation in smoking inequalities during the COVID-19 pandemic (42).

This study had a number of strengths. The sample size was orders of magnitude larger than any other study of smoking and COVID-19 and a broad range of data were collected, permitting the first analysis of smoking and COVID-19 infection in the population with adjustment for important confounding variables. The collection of data in real time while the pandemic is at its (anticipated) peak is also an advantage, minimising recall bias that is likely to be present in future studies that collect data retrospectively. There were also several limitations. First, rates of COVID-19 testing are lower in the UK than many other countries, suppressing numbers of confirmed COVID-19 cases; but this would only affect our results if testing rates differed by smoking status. Secondly, recency of cessation among former smokers was not assessed, so this group ranges from those who stopped smoking many years or decades previously to those who quit in the days or weeks prior to the survey. Future studies should ask former smokers about time since quitting in order to better evaluate whether abrupt quitting following the onset of COVID-19 symptoms may contribute to the lower rates of smoking recorded among hospitalised patients (4,16,17). Thirdly, the measure of smoking less underestimates those making reductions in their smoking because those who quit altogether in the last week are not included. Fourthly, the measure of changes in smoking was not anchored to the pandemic (i.e. did not ask about changes since COVID-19 started to affect the respondent’s life), so may not be sensitive to detect early changes in smoking in response to the pandemic. We observed associations between the date participants completed the survey and the odds of smoking less (which decreased over time) and smoking more (increased over time), which suggests changes in smoking behaviour in reaction to the pandemic may be changing as time passes, possibly in response to conflicting messages on smoking increasing or decreasing COVID-19 risk (27,28). Longitudinal data tracking changes over time within individuals would be useful in determining trajectories of smoking behaviour as the pandemic continues, and evaluating the extent to which any initial changes in smoking behaviour are maintained over time. Finally, while the sample was weighted to be representative of the UK adult population, results cannot be presumed to generalise to other countries with different demographic profiles or healthcare systems.

## Conclusions

When assessed by self-report in a population sample, current smoking was independently associated with increased odds of confirmed COVID-19 infection. There were socioeconomic disparities, with the association only apparent among those without post-16 qualifications. Smokers reported lower adherence to guidelines despite being more worried than non-smokers about catching or becoming seriously ill from COVID-19. Many smokers reported smoking more than usual, and COVID-19 related stress was associated with increased smoking, particularly among those with no post-16 qualifications.

There is a need to further investigate the unexpectedly low rates of smoking among hospitalised COVID-19 patients in order to establish why very different patterns of infection are being observed across smokers and non-smokers in clinical versus population settings. Any evidence of apparent protective effects of smoking should be interpreted cautiously with an awareness that it could encourage a surge in initiation of or relapse to smoking among never/ex-smokers looking to reduce their risk of COVID-19, the negative public health impact of which could far outweigh any purported benefit for COVID-19 outcomes.

## Data Availability

Anonymous data will be made available following the end of the UK pandemic.

## Declarations

### Ethics approval and consent to participate

Ethical approval for the COVID-19 Social Study was granted by the UCL Ethics Committee. All participants provided fully informed consent. The study is GDPR compliant.

### Transparency statement

SEJ affirms that the manuscript is an honest, accurate, and transparent account of the study being reported; that no important aspects of the study have been omitted; and that any discrepancies from the study as originally planned (and registered) have been explained.

### Availability of data and materials

Anonymous data will be made available following the end of the UK pandemic.

### Competing interests

JB has received unrestricted research funding from Pfizer, who manufacture smoking cessation medications. LS has received honoraria for talks, an unrestricted research grant and travel expenses to attend meetings and workshops from Pfizer, and has acted as paid reviewer for grant awarding bodies and as a paid consultant for health care companies. RW undertakes research and consultancy for and receives travel funds and hospitality from manufacturers of smoking cessation medications (Pfizer, GlaxoSmithKline and Johnson and Johnson). All authors declare no financial links with tobacco companies or e-cigarette manufacturers or their representatives.

### Funding

The COVID-19 Social Study was funded by the Nuffield Foundation [WEL/FR-000022583], but the views expressed are those of the authors and not necessarily the Foundation. The study was also supported by the MARCH Mental Health Network funded by the Cross-Disciplinary Mental Health Network Plus initiative supported by UK Research and Innovation [ES/S002588/1]. SJ and JB’s salaries were supported by Cancer Research UK (C1417/A22962). DF’s salary was supported by the Wellcome Trust [205407/Z/16/Z], The researchers are grateful for the support of a number of organisations with their recruitment efforts including: the UKRI Mental Health Networks, Find Out Now, UCL BioResource, HealthWise Wales, SEO Works, FieldworkHub, and Optimal Workshop. JB, LS and SJ are members of SPECTRUM a UK Prevention Research Partnership Consortium [MR/S037519/1]. UKPRP is an initiative funded by the UK Research and Innovation Councils, the Department of Health and Social Care (England) and the UK devolved administrations, and leading health research charities. The funders had no final role in the study design; in the collection, analysis and interpretation of data; in the writing of the report; or in the decision to submit the paper for publication. All researchers listed as authors are independent from the funders and all final decisions about the research were taken by the investigators and were unrestricted. All authors had full access to all of the data (including statistical reports and tables) in the study and can take responsibility for the integrity of the data and the accuracy of the data analysis.

### Authors’ contributions

SEJ, LS, DF, AS, and JB conceived and designed the study. SEJ analysed the data and wrote the first draft. LS, DF, AS, and JB provided critical revisions. All authors read and approved the submitted manuscript.

## Notes

### Clinical Protocols

https://osf.io/pcs49/

